# WAVELET-BASED AUTOMATIC PECTORAL MUSCLE SEGMENTATION FOR MAMMOGRAMS

**DOI:** 10.1101/2024.02.09.24302580

**Authors:** Basel White, Amy Harrow, Christina Cinelli, Kendra Batchelder, Andre Khalil

## Abstract

The computational analysis to assist radiologists in the interpretation of mammograms usually requires a pre-processing step where the image is converted into a black and white mask to separate breast tissue from the pectoral muscle and the image background. The manual delineation of the breast tissue from the mammogram image is subjective and time-consuming. The 2D Wavelet Transform Modulus Maxima (WTMM) segmentation method, a powerful and versatile multi-scale edge detection approach, is adapted and presented as a novel automated breast tissue segmentation method. The algorithm computes the local maxima of the modulus of the continuous Gaussian wavelet transform to produce candidate edge detection lines called maxima chains. These maxima chains from multiple wavelet scales are optimally sorted to produce a breast tissue segmentation mask. The mammographic mask is quantitatively compared to a manual delineation using the Dice-Sorenson Coefficient (DSC). The adaptation of the 2D WTMM segmentation method produces a median DSC of 0.9763 on 1042 mediolateral oblique (MLO) 2D Full Field Digital mammographic views from 82 patients obtained from the MaineHealth Biobank (Scarborough, Maine, USA). Our proposed approach is evaluated against *OpenBreast*, an open-source automated analysis software in MATLAB, through comparing each approach’s masks to the manual delineations. *OpenBreast* produces a lower median DSC of 0.9710. To determine statistical significance, the analysis is restricted to 82 mammograms (one randomly chosen per patient), which yields DSC medians of 0.9756 for the WTMM approach vs. 0.9698 for *OpenBreast* (*p*-value = 0.0067 using a paired Wilcoxon Rank Sum test). Thus, the 2D WTMM segmentation method can reliably delineate the pectoral muscle and produce an accurate segmentation of whole breast tissue in mammograms.

## INTRODUCTION

Breast cancer is the most common cancer among women, affecting one in eight [1]. The introduction of computer-aided detection (CADe), diagnosis (CADx), and triage (CADt) have rapidly become useful tools in clinical support systems for screening mammography and the use of CAD in a clinical setting has increased from 5% in 2003 to 92.3% in 2016 [2]. CADx and CADe allow radiologists to augment their visual assessment by marking suspected abnormalities and providing the probability of malignancy in the cases of CADe., while CADt systems are designed to prioritize suspicious studies [3]. Unfortunately, current CAD methods have been associated with decreased specificity, increased recall rates of healthy women, and false positives on up to 70% of normal cases of breast cancer [4-6]. More recently, deep learning models have shown the potential to improve detection rates, lower recall rates, and assess future cancer risk. However, their predictive power may be diminished for extremely dense breasts, which are women who are also at the greatest risk of cancer [7, 8]. In addition to the limitations of deep learning approaches due to their inexplicability [9], they may be strongly affected by otherwise inconsequential routine equipment maintenance. For example, one study showed that recall rates increased approximately threefold following a software upgrade on the mammography equipment [10]. Taken with the high computational costs of deep learning approaches, interpretable methods are being developed, such as our patented method that is not based on deep learning [11, 12] and is designed to assess the breast tissue microenvironment from mammograms through the use of a wavelet-based multifractal image analysis formalism [13, 14], a core part of the proposed approach in this manuscript.

Prior to performing a computational analysis on a mammogram, a spatial representation (i.e. a mask) of the breast tissue is typically required. A common, yet subjective, method for creating a mammographic mask is for a person to manually trace the outline of the breast. Manual segmentation is tedious due to the click-and-trace nature of following along the pectoral muscle contour and the rest of the breast from the image background. The sheer nature of human variability regarding the shape of the breast, pectoral muscle, and, through the technician imaging the breast causes the segmentation of the pectoral muscle from mammograms to be an innate problem in the creation of these masks. Such subjectivity and variability can influence the subsequent computational analysis of the mammograms. An automatic method allows for the limitations associated with the manual masking process and the human subjectivity associated with delineating the breast tissue to be eliminated.

The pectoral muscle area of the mammogram is usually high in pixel intensity, and similar to the intensity seen in that of dense breast tissue. The similarity in the intensity of the dense breast tissue and the pectoral muscle area causes failures for off-the-shelf automated threshold-based segmentation techniques, based on our early trials [data not shown]. Yet there is a gradient lying at the boundary between breast tissue and the pectoral muscle region of the breast (**Figure 1a**). A wavelet-based pectoral muscle segmentation algorithm for grayscale mammograms was constructed through the adaptation of the 2D Wavelet-Transform Modulus Maxima (2D WTMM) segmentation method. Using multi-scale edge detection lines, the 2D WTMM segmentation method can be used to perform automatic image segmentation for a variety of images across scientific fields. Indeed, the 2D WTMM segmentation method was first developed and used for the automatic segmentation and analysis of the morphology of interphase chromosome territories from fluorescence microscopy images [15]. Additionally, this method was implemented for the automatic detection of aggregates in ultra-thin gold surfaces [16], coronal loops in ultraviolet (UV) images of the solar corona [17], solar photospheric magnetic structures [18], and C. elegans embryonic cell nuclei in three-dimensional image stacks from fluorescence microscopy [19]. The 2D WTMM segmentation method was also implemented to detect and characterize clusters of microcalcifications from mammograms [20-22], track bacteria in digital holographic microscopy time series [23], as well as for tracking glacier termini from satellite imagery in southeast Greenland [24].

**Figure 1:**
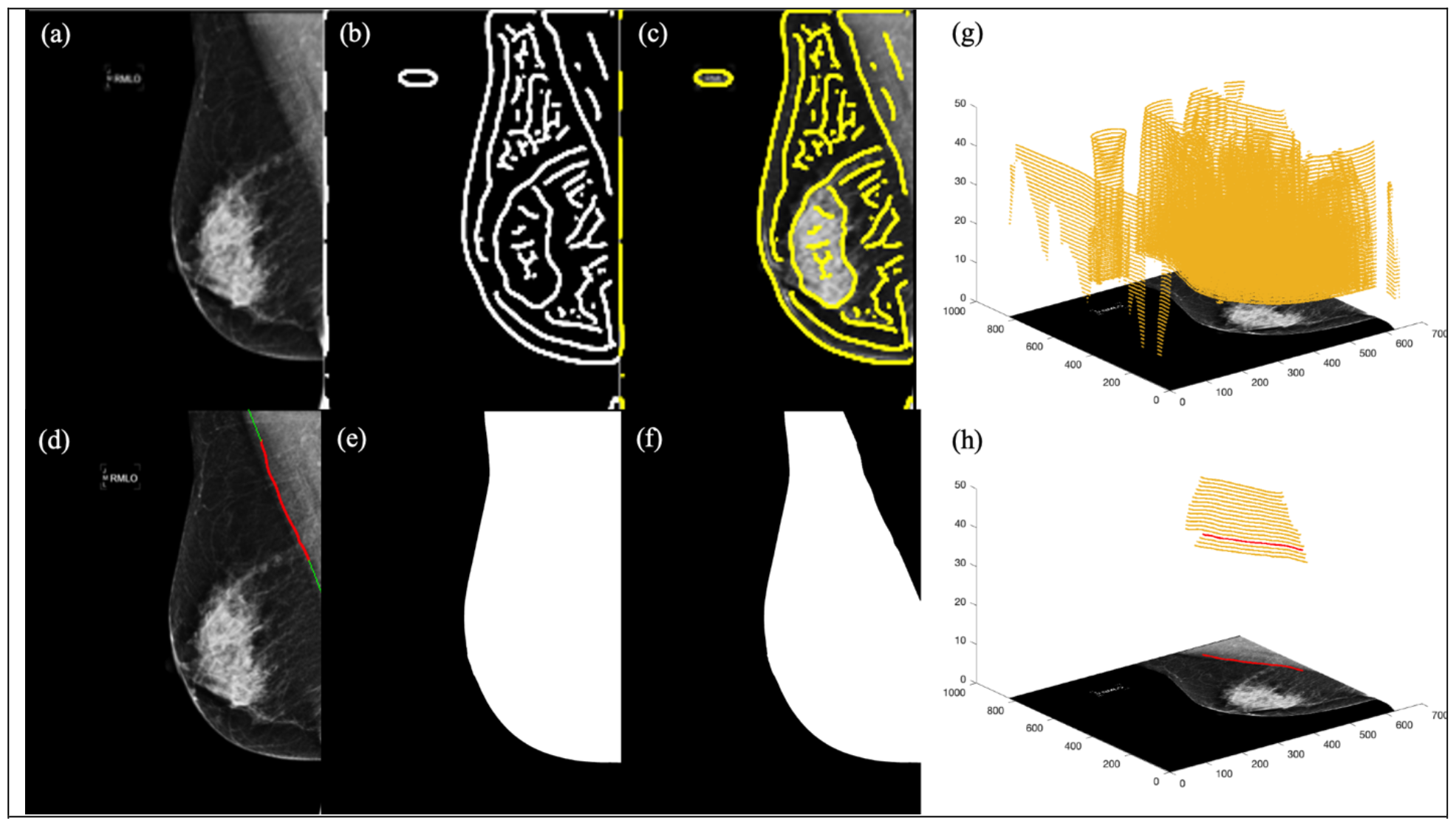
For a sample MLO mammographic view (a), the WTMM chains at scale 30 (b) are overlaid onto the mammogram (c). The final pectoral muscle chain (red) and linear chain extensions (green) are overlaid onto the mammogram (d). The segmented pectoral muscle region can be subtracted from the whole thresholded MLO mammogram (e) to create the final automatic binary mask from the 2D WTMM segmentation method (f). Maxima chains across 50 wavelet scales for sample MLO mammogram are shown in (g) and the candidate chains (yellow) and final pectoral muscle maxima chain (red) are shown in (h).

### Related Works

Optimized thresholding can be utilized to acquire the most efficient threshold to segment only the pectoral muscle region of the grayscale mammogram rather than breast tissue [25, 26]. Challenging issues with these approaches occur when similar pixel intensities are found between the pectoral muscle region and dense breast tissue. Another limiting factor may be related to the assumption that the pectoral muscle region must be triangular. The similar pixel intensities from these two regions can be utilized in a region-growing approach in which a uniform intensity value (UIV) is calculated based upon the mean and standard deviation of pixel values, excluding those equal to zero, across the grayscale mammogram [27]. The UIV can then be used to fully segment the pectoral muscle region, in which curve fitting is included as the final step to insure the entire pectoral muscle region has been segmented. In a different approach, the pectoral muscle boundary is acquired through the use of thresholding, in which an active contour model can be used to search for the true pectoral muscle contour [28]. A graph-cut technique can be utilized to create a region of interest (ROI) containing the entire pectoral muscle region of the breast [29]. The similar intensities present in this region are used to ensure the entire pectoral muscle region is acquired in this ROI with the help of Bezier curves, used widely in computer graphics to represent smooth curves. The top right corner of the ROI can then be selected as a control point for the Bezier curve in which an iterative method can be implemented for the final construction of a smoother Bezier curve representing the pectoral muscle contour. Genetic algorithms, morphological methods, and random sample consensus approaches have also been explored [30, 31].

Machine learning methodologies are also utilized for the segmentation of the pectoral muscle from mammograms [26, 32-34]. An example of this is seen through a connected component labeling method to remove the pectoral muscle region of the breast from the mammogram. All mammograms are flipped to the left MLO orientation in which Otsu’s multi-thresholding approach is used to separate the background, high, and low dense regions of the respective input mammogram [26]. The company Volpara™designed an implementation of the *U*-net machine learning algorithm [35] to segment both the breast and pectoral region of the mammogram. Volpara’s™ machine learning approach additionally utilizes image normalization, algorithmic padding, image sizing and contrast adjustment, and altering image resolution to improve algorithm efficiency [33]. Fuzzy C-Means clustering algorithms can be utilized to acquire the pectoral muscle region of the breast, while the final pectoral muscle contour is acquired through iterative contour improvement and validation [32].

An open-sourced MATLAB software, *OpenBreast*, employs an automated breast and chest wall segmentation algorithm in their breast cancer risk evaluation pipeline [36]. For breast segmentation, *OpenBreast* employs an established thresholding methodology in which the threshold is determined based upon the highest value of the distribution of pixel intensities of the mammogram [36]. Once the breast is segmented, the chest wall is then segmented through the use of the Hough-based edge detector methodology [36]. The *OpenBreast* algorithm was tested on a total of 305 2D full-field digital mammography images from the Breast Cancer Digital Repository [36].

The use of deep and machine learning algorithms seems to perform well in segmenting the pectoral muscle. However, weaknesses seen through these literature reviews lie in the assumption of the pectoral muscle as a straight line, which indicates breast tissue loss in the creation of binary masks, which creates an opportunity for a missed cancer. Furthermore, the necessity to train the respective machine learning, and/or the inability to explain the model’s predictions present limitations. The training process takes time and can be swayed due to poor model construction or even the presence of an unbalanced training sample set. **Table 1** reports the performances of the approaches described above in which varying datasets were used.

**Table 1:** Metrics used in literature to evaluate current methodologies in pectoral muscle segmentation.

| Study | Evaluation Metrics* | Mean Results |
| --- | --- | --- |
| Rahimeto et al. 2021 [26] | Accuracy and IoU | 0.9862 and 0.8362 |
| Gomez et al. 2021 [27] | Accuracy | 0.95 |
| Rampun et al. 2017 [28] | DSC | $0.963 \pm 0.026$ |
| Camilus et al. 2009 [29] | FN and FP | 0.0558 and 0.0064 |
| Shen et al. 2018 [30] | FN and FP | Three datasets: (1) 0.0203 and 0.0690; (2) 0.0160 and 0.0403; (3) 0.0242 and 0.1361 |
| Feudijo et al. 2013 [32] | FN and FP | $0.1112 \pm 0.1253$ and $0.0335 \pm 0.872$ |
| Wang et al. 2019 [33] | DSC | 0.8879 |
| Yu et al. 2022 [34] | IoU and DSC | $0.9746 \pm 0.0045$ and $0.9630 \pm 0.0066$ |
| Yoon et al. 2016 [31] | Accuracy | 0.922 |

\*IoU: Intersection over Union; DSC: Dice-Sorenson Coefficient; FN: False Negative Rate; FP: False Positive Rate

## MATERIALS AND METHODS

### The 2D Wavelet Transform Modulus Maxima Segmentation Method

The 2D WTMM segmentation method is a multi-scale, gradient-based method that identifies contours representing the locally maximal changes in intensity in an image. A 2D smoothing Gaussian function is used, denoted as 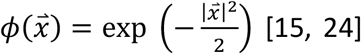 [15, 24], where 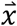 represents a coordinate (*x*_1_, *x*_2_) and 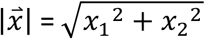. The continuous wavelet transform is then calculated by the partial derivatives of this smoothing function 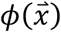 with respect to *x*_1_ and *x*_2_, respectively, 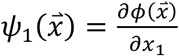 and 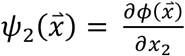. The gradient is calculated through convolving the image with the 2D smoothing function, where * represents the convolution in the image, 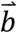 represents the position, and *a* represents the scale of the convolution: 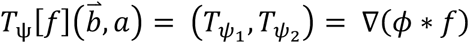. The wavelet transform is performed at 50 different size scales, *a* = 0, 1, …, 49, where the size scale in pixels is 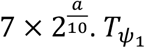 and 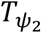 represent the two components of the wavelet transform:

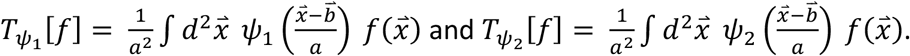

The resulting gradient is in vector form, meaning it has both a magnitude and direction. The magnitude of the gradient is known as the wavelet transform modulus *M*_ψ_, while the direction is known as the argument *A*_*ψ*_ pointing to the largest intensity variation in an image:

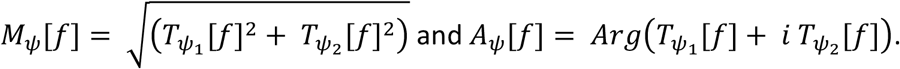

The maxima points, known as the wavelet transform modulus maxima, or WTMM, represent the positions in the image where *M*_ψ_ are maximal. These maxima points are organized into maxima chains, which can be utilized as edge detection lines (**Figure 1**).

### Dataset

Our algorithm was developed on a dataset of 1042 de-identified 2D digital full field “FOR PRESENTATION” mammograms from the MaineHealth Biobank, Scarborough, Maine. These mammograms are all MLO views at 70 microns per pixel (vendor = Hologic) and were obtained under MaineHealth IRB approval #4664 in 2015. The same mammograms were used to evaluate the performance of the 2D WTMM segmentation method and the open-source *OpenBreast* approach when compared to manual delineations.

### Radiological Assessment of Mammographic Density

Mammographic breast density is quantified by radiologists using the Breast Imaging and Reporting Data System (BI-RADS) recommendations from the American College of Radiology. In its 5^th^ edition, the BI-RADS density levels are defined as A: Almost entirely fatty; B: Scattered fibroglandular densities; C: Heterogenously dense; and D: Extremely dense [40]. For each of the 82 patients, breast density was visually assessed by two breast radiologists (AH and CC). Each patient’s mammographic density was categorized as either non-dense (BI-RADS A or B) or dense (BI-RADS C or D).

### Manual Delineations

To create ground truth masks, all 1042 mammograms were individually loaded into the image analysis software FIJI [39]. The polygon selection tool was used to manually select the region of breast tissue, excluding the pectoral muscle area. Aber clearing the outside of the polygon, the resulting image was saved as a black and white binary mask. A subset of 217 MLO mammograms (∼20% of the dataset) were manually delineated by two different people. This was to allow us to perform an inter-human variability analysis and empirically determine the maximal achievable DSC score when comparing the automated methods to manual delineations. To evaluate the similarity between two human delineators, we designed an inter-human variability (IHV) analysis. The IHV analysis helps determine a “ground truth” DSC to which an automated masking process can be compared to.

### WTMM Breast Segmentation Approach

As a preprocessing step, the MLO mammogram is scaled down by a factor of 4 in the *x* and *y* directions using a pixel averaging algorithm allowing for faster processing without compromising accuracy. The 2D WTMM segmentation method is then called, and the scale-by-scale maxima chain information is saved to file for all 50 wavelet scales. At each scale, the descriptive metrics of every WTMM chains are recorded: *size, mean-modulus, mean-argument, stdev-argument, mass, posfirst*, and *distance*, where *size* represents the number of maxima points constructed into the chain (i.e. its length); *mean-modulus* describes the average modulus value of the maxima chain; *mean-argument* and *stdev-argument* describes the average and standard deviation, respectively, of the argument value for the maxima chain; *mass* is the product of size and mean-modulus; *posfirst* is the position of the first point on the maxima chain (used as a coordinate reference); and *distance* is the Euclidean distance between the geometrical center of the maxima chain and the top left or right corner of the mammogram, depending if the view is from the left or right breast.

Empirically determined thresholds were based upon calibrated average values of these descriptive metrics. The implementation of these thresholds allows for candidate maxima chains to be selected (**Figure 1h**). Of these candidate maxima chains, the maxima chain with the lowest *stdev-argument* is deemed as the maxima chain most accurately matching the pectoral muscle contour, as shown in red in **Figures 1d and 1h**. In some cases, the final pectoral muscle maxima chain does not extend to the top and side of the image, where a linear extension is added based on the linear regression fit of the final pectoral muscle maxima chain as shown in green in **Figure 1d**. Once the final pectoral muscle maxima chain is acquired, the final step of the algorithm is the construction of the binary mask by combining the segmented pectoral muscle region with an automated intensity-based segmentation of the whole breast.

### Dice-Sorenson Overlap Coefficient

For two binary mask image ROIs, *X* and *Y*, where *X* is the manually delineated mask and *Y* is the automated mask obtained by either the WTMM or *OpenBreast* approaches, the Dice-Sorenson Coefficient (DSC) is defined as the ratio of twice the area of intersection between the two ROIs over the sum of the individual ROI areas [38], i.e. 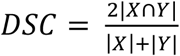. The DSC ranges from 0 to 1, where a value of 1 indicates a perfect overlap between the two ROIs.

### Statistical Analysis

All statistical distribution analyses, and hypothesis testing yielding the *p*-values presented in this article were performed using the R language, version 4.3.1 [37].

### RESULTS

Segmented mammograms from the automatic 2D WTMM segmentation algorithm were quantitatively compared to manually drawn masks via the DSC on a total of 1042 MLO mammograms resulting in a median DSC of 0.976 (**Figure 2**). To evaluate the performance of the 2D WTMM segmentation method, *OpenBreast* was employed on the 1042 MLO mammograms from the MaineHealth mammographic dataset and compared to their manually drawn masks resulting in a median DSC of 0.971. To determine statistical significance, the analysis was restricted to 82 mammograms (one randomly chosen per patient), which yielded DSC medians of 0.9756 for the WTMM approach vs. 0.9698 for *OpenBreast* (*p*-value = 0.0067 using a paired Wilcoxon Rank Sum test). We also assessed the potential differences in the masking performance of the automated methods with different mammographic breast densities. The 82 patients were categorized as non-dense (*n*=47) if their BI-RADS mammographic density, as visually assessed by radiologists (AH and CC), was either A or B, or dense (*n*=35) if their BI-RADS mammographic density was either C or D. The performances were not statistically different when comparing the non-dense vs dense subgroups for OpenBreast (*p-value* = 0.4282 using a Wilcoxon Rank Sum test) or the WTMM (p-value = 0.3936 using a Wilcoxon Rank Sum test). A median DSC for the IVH analysis for the subset of 217 MLO mammograms from these analyses was found to be 0.983.

**Figure 2:**
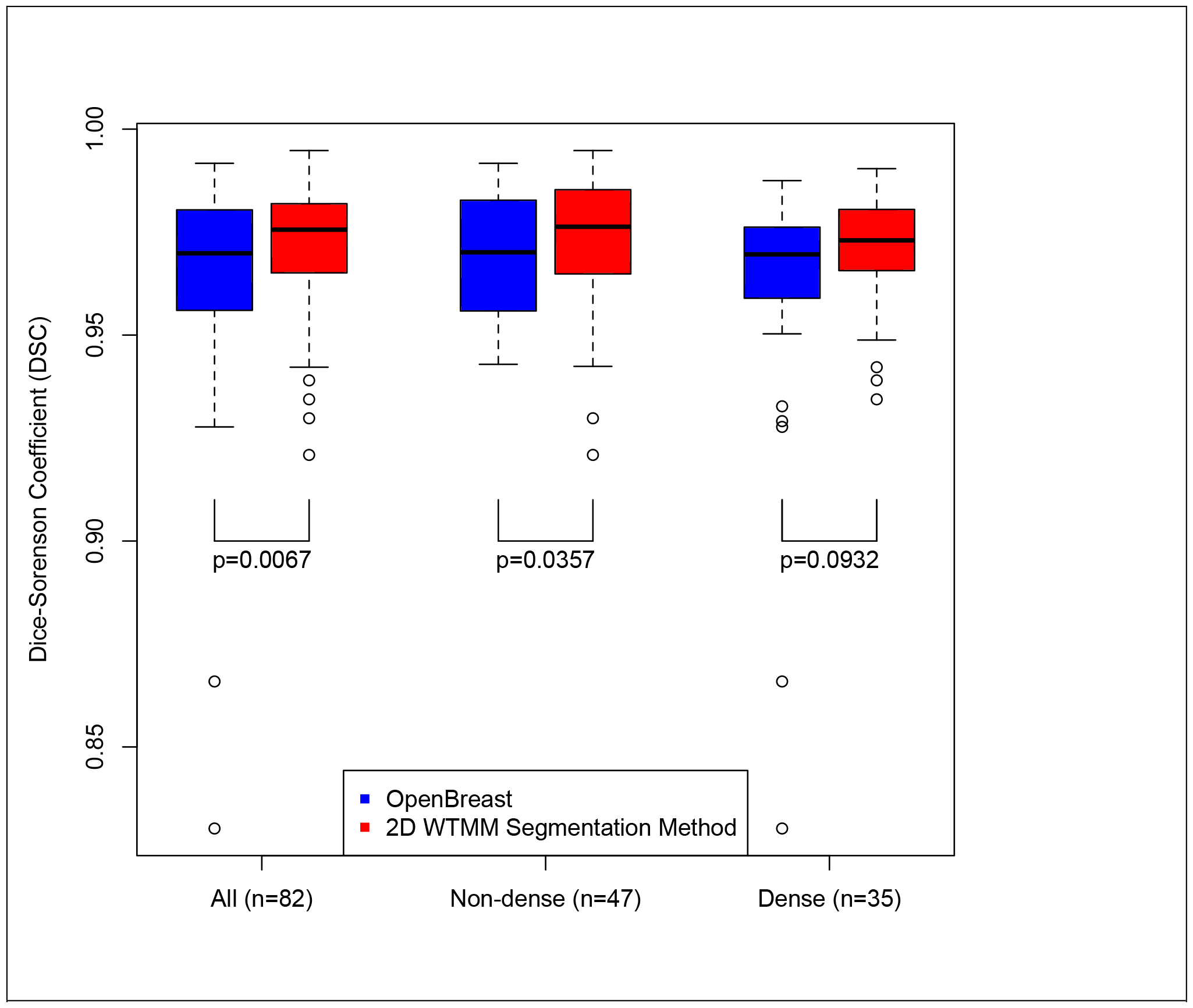
The median DSCs are compared for the 2D WTMM segmentation method (red) and *OpenBreast* (blue).

**Figure 3:**
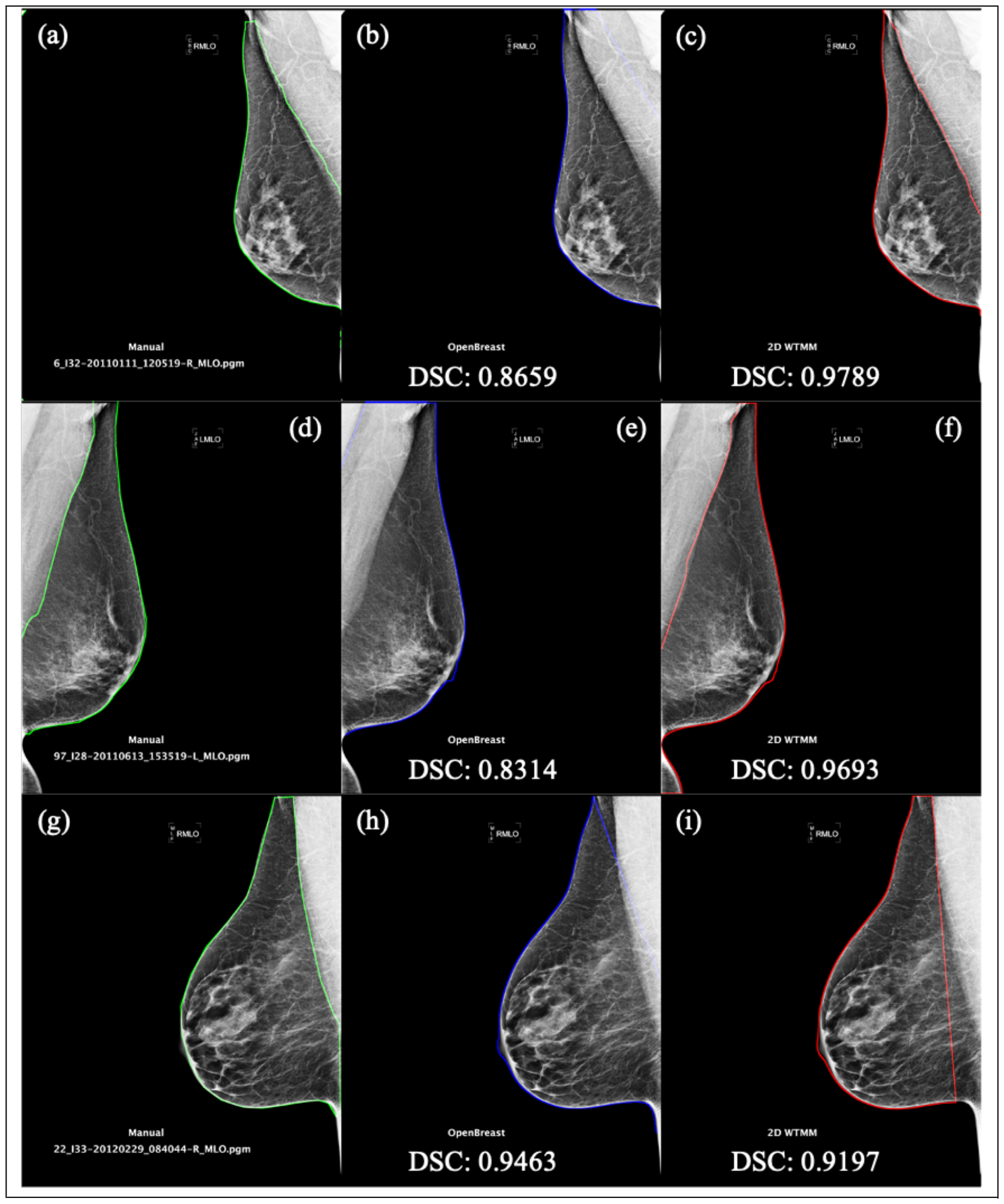
Three sample mammograms overlaid with their human delineations (a)(d)(g), *OpenBreast* (b)(e)(h), and 2D WTMM segmentation method (c)(f)(i).

## DISCUSSION

The segmentation of the pectoral muscle and image background from mammograms is a necessary preprocessing task for various computational analyses. Manual delineation from visual inspection is subjective, time-consuming, and does not integrate well into an automated analysis workflow of large sets of mammograms, while potentially reducing the computational expense of machine learning approaches. We propose a novel automated method to segment the pectoral muscle in MLO mammographic views using the 2D WTMM segmentation method. The performance of our wavelet-based breast segmentation analysis produced a higher median DSC in comparison with the *OpenBreast* analysis for MLO mammograms. With this, the 2D WTMM segmentation method is an efficient and automatic segmentation approach for mammograms, yielding a high accuracy. Breast density is one of the strongest risk factors for breast cancer [41] with an almost five-fold relative risk for extremely dense breasts compared to fatty breasts [42]. The performance of both automated methods did not seem to be affected by the mammographic density of the breast being masked. However, the results are suggestive that the WTMM is more effective in patients with dense breasts than *OpenBreast*. Further development of the WTMM approach includes additional validation on an even larger dataset, with a diversity of mammography manufacturers, and from a diverse patient population. In addition to this, the WTMM approach is in the process of being adapted into an open-source Python approach.

## Data Availability

The authors will make the data available upon reasonable request.

## DATA AVAILABILITY STATEMENT

The authors will make the data available upon reasonable request.

## AUTHOR CONTRIBUTIONS

BW, AK: WTMM and *OpenBreast* analysis. KB: statistical analyses. BW, KB, AK: figure preparation and manuscript writing. AH and CC: Radiological breast density assessments. All authors have read and approved of the manuscript.

## ACKNOWLEDGEMENTS

We are grateful to Drs Anne Breggia, Ivette Emery, and Joe Schulte from MaineHealth for the mammography database, and to CompuMAINE Lab members Arihant Tallapureddy, Melissa Ham, and Sarah Glattr for the manual delineations. We also thank Drs. Karissa Tilbury and Peter Stechlinski for technical discussions.

